# Age Bias, Missing Data, and Declining Response Rates in the National Youth Risk Behavior Survey and Their Influence on Estimates of Trends in Adolescent Sexual Experience, 2011-2023

**DOI:** 10.1101/2025.03.05.25323024

**Authors:** John J. Santelli, Debbie Malden, Regan A. Moss, Mikal Finkelstein, Laura D. Lindberg

**Author notes:** **Corresponding author:** John Santelli, 722 West 168th Street New York, NY 10032, United States.

## Abstract

**Introduction:** The national Youth Risk Behavior Survey (YRBS) has experienced considerable declines in response rates, increases in missing data on sexual experience, and shifts in data collection – all of which raise questions about bias in estimates for sexual experience among US high school students.

**Methods:** We used weighted data from the YRBS for 2011-2023 (n=110,409). We explored the impact of declining school and student response rates, missing data, and shifts in age distribution in 2021 on reported sexual intercourse. Using statistical decomposition, we estimated the percentage change in this outcome between 2019 and 2021 due to change in the age structure versus changes in reported behavior.

**Results:** From 2011 and 2023, school and student survey response rates declined (school 81% to 40%, student 87% to 71%, and overall, 71% to 35%). Missing data on ever had sex increased over time from 7.0% in 2011 to 29.5% in 2019 and 19.8% in 2023. The age structure in the YRBS national sample was similar from 2011-2019 and in 2023, but substantially younger in 2021. Statistical decomposition estimated that 50% of the change in sexual experience among adolescent women between 2019 and 2021 and 30% of the change for adolescent men was due to a change in the age distribution.

**Implications:** Declining response rates, increased missing data, and changes in the age structure of 2021 YRBS raise serious concerns about the validity of trends in the YRBS. A concerted national effort is needed to build support for the collection of YRBS and other public health surveillance data.

## Introduction

Monitoring trends in adolescents’ sexual behavior is essential to understand social and reproductive health outcomes and the impact of public policies and programs and other experiences on young people’s health and well-being. Adolescence is a period of sexual exploration for many; estimates from United States from 2015-2019 find that the share of female adolescents who have ever had sex rises from 3% at age 13 and 10% at age 14, to 64% at age 18 and to 71% at age 19. Similar patterns by single year of age are found among male adolescents^1^. Considerable diversity in the timing of sexual initiation among nations is found - over time and often between the sexes^2,3^.

The Centers for Disease Control and Prevention (CDC) has reported sharp declines in many adolescent health behaviors including sexual intercourse between 2019 to 2021 among high school student in the national Youth Risk Behavior Survey (YRBS)^4^.This two-year period is of particular interest as it includes the Covid-19 pandemic which brought about many changes in adolescents’ lives, including school closures, that may have impacted adolescents’ sexual behaviors^5^. While the behavioral changes measured between 2019-2021 may be real, change in the timing of YRBS data collection in 2021 may have influenced reported estimates of adolescent behaviors. The YRBS has historically been administered in the spring thru 2019, but in 2021 the YRBS was administered in the fall, due to COVID-19. While the target population remains students in grades 9 – 12 in each year, high school students are systematically younger in the fall compared to the spring and may differ on other parameters, raising concern that the 2021 YRBS is not a comparable sample to earlier survey years.

Likewise, it has become increasingly difficult to collect data in US national surveys including the YRBS; multiple federal surveys have reported sharply declining response rates^6^. The reasons for these declines is poorly understood^6^. States may refuse to participate in the national and state YRBS^7,8^; even when states participate in the YRBS, response rates at both the school-level and student/parent-level have declined over time9. Additionally, among participating states, there has been increasing refusal to include questions on sexual and reproductive health^10^.

Accurate measurement is essential in national public health surveillance^11^. Accuracy in surveys with adolescents is dependent on multiple factors, such as how data are collected (face-to-face, telephone, paper and pencil or on computer), adolescent perceptions of confidentiality, the clarity of question wording, the use of parental permission, response bias, and the precision of sampling and recruitment^12^. Scientists who use population-based data are careful to use consistent methods over time and to estimate the validity and reliability of surveillance systems. For example, the YRBS publishes regular methodological reports^9^, generally uses consistent question wording, and periodically reports on reliability of the questions in the YRBS [13, 14]. The YRBS has also examined the impact of written parent permission vs. so-called passive parental consent^15^. Overall, the CDC has labored to be consistent in YRBS measurement and YRBS has demonstrated to be reliable when tested^13,14^.

In this study, we initially had planned to explore changes in the age structure of the national YRBS sample in 2021 and the impact of age structure on trends from 2011 to 2021 in the prevalence of sexual experience (reporting ever had sexual intercourse). Anomalously in 2021, the YRBS was administered in the fall of the school year given the COVID pandemic. We then became aware of increases in missing data and declining response rates; the later has been reported by CDC^16^. We decided it was important to explore all three issues to provide a more complete picture of methodological challenges in the national YRBS in the measurement of sexual experience.

## Methods

### Data sources

This study used secondary data from the national YRBS, a population-representative school-based survey of high school students conducted biennially by the Centers for Disease Control and Prevention (CDC). The YRBS is designed to measure the prevalence of various health behaviors, including sexual activity, among population-representative samples of public and private school students in 9^th^ through 12^th^ grades in the United States^10^. The survey is self-administered anonymously in a computer-scannable booklet (i.e., paper and pencil administration) and takes one class period (approximately 45 minutes) to complete. CDC’s Institutional Review Board approved the protocol for the YRBS. For this analysis, we used publicly available data and thus were exempt from further IRB review.

The Youth Risk Behavior Surveillance System (YRBSS) is the largest public health surveillance system in the United States, monitoring a broad range of health-related behaviors among high school students^9^. The system includes a nationally representative survey, as well as representative surveys of some states, tribes, territories, and local school districts; the sampling for state and national surveys may also overlap. YRBS uses a three-stage stratified clustered sampling and oversamples Black and Hispanic respondents; data are weighted based on the current population of high school students as reported by US DOE (Department of Education). The three stages include school systems, schools, and classrooms within schools. School systems are selected based on population size. Weights for the biennial data from the YRBS produce nationally representative samples of high school students for each survey year. Weighted data account for the distribution of the sample by grade but not age. Additional information about YRBS is available online (www.cdc.gov/healthyyouth/data/yrbs/) and in methodological reports^9^.

States may refuse to participate in the state YRBS and in the national YRBS. Additionally, school systems, individual schools, parents, and adolescents may refuse to participate. State participation in state-based YRBS varied by survey year, but it is unclear how this influences the national sample.

Both parental permission and adolescent assent are required for participation. Parents/ guardians provide either active (signed) permission or passive consent (a letter to parents with the potential for refusal)^15^. Non-response rates in the national YRBS are reported as: 1- schools refusing to participate and 2- students refusing to participate, including refusal by either parent/ guardian or student.

Since its inception, the YRBS has been conducted during the spring (January-June). However, due to the impact of the COVID-19 pandemic on school attendance through school closures and instructional models, the 2021 YRBS was postponed until fall (September-December) 2021^9^, but no direct age adjustment for this change in timing was made. The 2021 data were weighted by grade but not by age.

### Measures

The variables in these analyses were limited to age, gender (female and male students) and ever had sex based on the questions “have you ever had sexual intercourse?” Given missing data on ever had sex variable, we also used the question on reported ‘age at first sex,’ which includes ‘never had sex’ as a response option.

### Analyses

Our analyses examined the period 2011-2023, with a specific focus on changes from 2019-2021. We described the age distribution (including median and mean age) of the sample by year and gender to examine the influence of the 2021 shift in the timing of the survey administration on students’ ages. Linear trends were assessed using linear regression and heterogeneity was assessed using chi-square statistic, and p-values of ≤0·05 considered statistically significant. For adolescents aged ‘14 and under’ and ‘18 and over’, the exact age was unknown. Hence, we imputed age to 1 decimal place within each age group by using multiple imputation with 20 repetitions, assuming the distribution in birth month did not change by year, allowing a small degree of uncertainty across age groups^17^. We estimated the proportion who reported ever having sex by gender in each year unadjusted and adjusted for age group. In adjusted analyses, we applied a direct standardization approach using the 2019 YRBS population as the reference population for age distribution^18,19^; the unadjusted rates match those previously published by CDC^9^. We calculated the differences between 2019 and 2021 in the unadjusted and adjusted rates of sexual experience and decomposed the change from 2019 to 2021 in the age-adjusted rates into the proportion due to shifts in the age composition versus shifts in adolescent reporting of ever had sex^18^.

Following CDC guidelines, data were weighted to adjust for nonresponse and oversampling of Black and Latinx youth. We started with the complete sample of 110,409 students. Analyses were restricted to participants with complete data on age, gender and ever had sex (*n* =93,046) for the primary analysis of ever had sex. The sensitivity analysis included 100,305 respondents, supplementing the primary study population with additional respondents who had missing information on ‘ever had sex’ but complete information on ‘age at first sex’. **Supplementary** Figure 1 shows the study population flow chart for the primary and sensitivity analyses. All analyses were conducted in Stata (version 18.0) and SAS (version 9.4). Visual representations were created using R (version 4.4.1).

## Results

Student and school response rates decreased over time (**Figure 1A**) - as has been reported by CDC [9] - From 2011 to 2023 school and student survey response rates (RR) declined considerably (school RR=81% to 40%, student RR=87% to 70%, and overall RR=71% to 35%). The sharpest declines occurred among schools and between 2021 and 2023.

**Figure 1.**
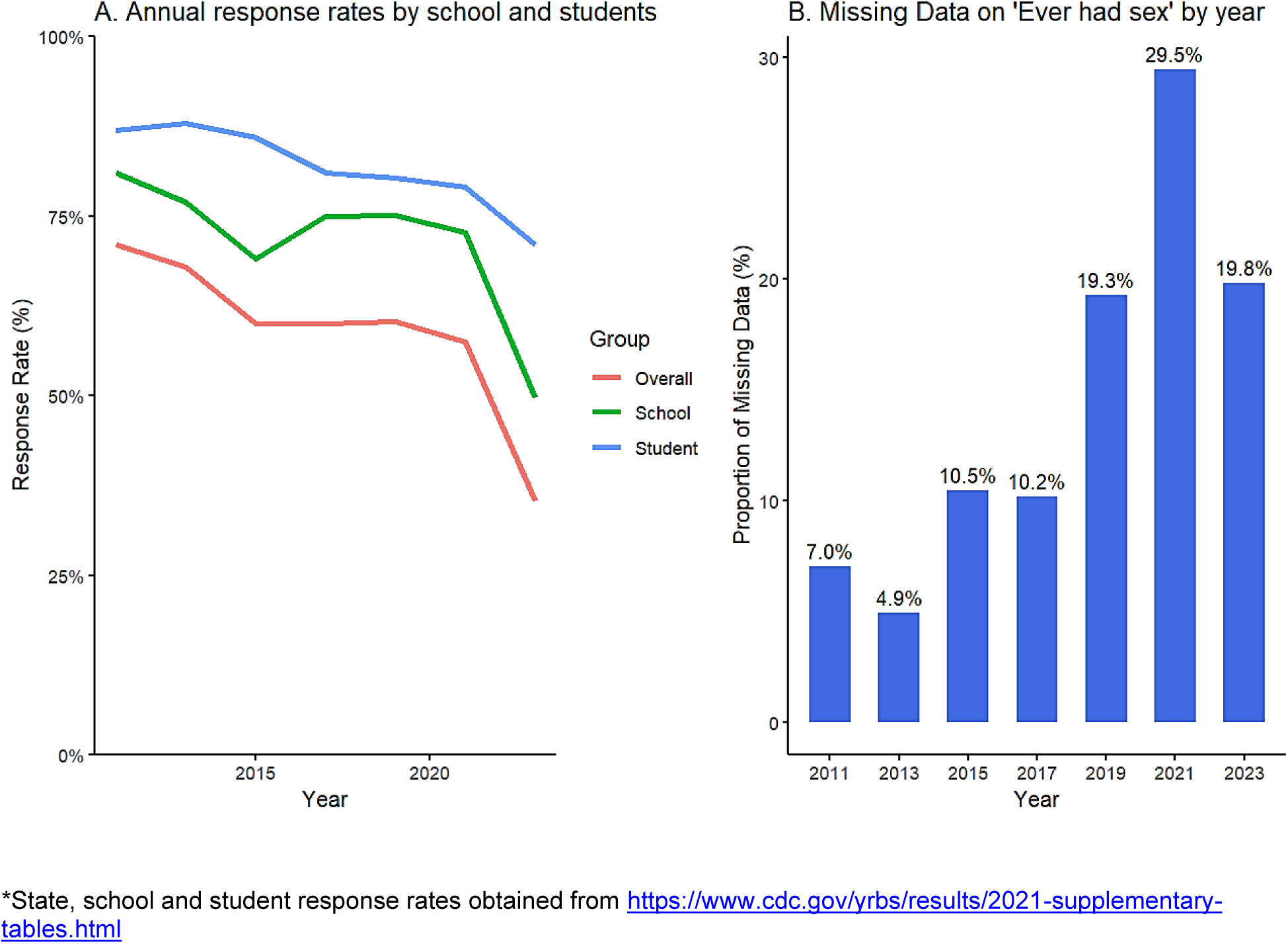
Survey response rates at the school- and student-level* and missing data on ‘ever had sex’ survey question, national YRBS, 2011-2023

Missing data on the ‘ever had sex’ question item increased sharply over time from 7.0% in 2011and 4.9% in 2013 to 29.5% in 2021 and 19.8% in 2023 (**Figure 1B)**, reflecting more schools not asking the ‘ever had sex’ question. In general, missing data on ever had sex was more common among younger students (data not shown). Among respondents, the national YRBS sample had smaller percentages of missing data on gender (0.7%, n=673), age (0.5%, n=456), or both (1.0%, n=896) than ever had sex; missing data on gender increased over time from 0.4% in 2011 to 1.4% in 2021 and then declined in 2023 (0.8%) [**Table 1**].

**Table 1.** Baseline characteristics of national YRBS survey participants using CDC defined weights*, by gender, age and study year, 2011-2023.

|  | Survey year |  |  |  |  |  |  |
| --- | --- | --- | --- | --- | --- | --- | --- |
|  | 2011, N (%) | 2013, N (%) | 2015, N (%) | 2017, N (%) | 2019, N (%) | 2021, N (%) | 2023, N (%) |
| **Total** | 15425 | 13583 | 15624 | 14765 | 13677 | 17232 | 19939 |
| **Gender** |  |  |  |  |  |  |  |
| Female adolescents | 7446 (48.3%) | 6780 (49.9%) | 7551 (48.3%) | 7428 (50.3%) | 6690 (48.9%) | 8201 (47.6%) | 9573 (48.0%) |
| Male adolescents | 7925 (51.4%) | 6790 (50%) | 7955 (50.9%) | 7224 (48.9%) | 6862 (50.2%) | 8783 (51.0%) | 10314 (51.7%) |
| Missing | 54 (0.4%) | 12 (0.1%) | 119 (0.8%) | 114 (0.8%) | 125 (0.9%) | 248 (1.4%) | 158 (0.8%) |
| **Male adolescents** |  |  |  |  |  |  |  |
| **Age, years<sup>§</sup>** |  |  |  |  |  |  |  |
| ≤14 | 803 (10.9%) | 639 (10.0%) | 646 (9.1%) | 723 (11.1%) | 677 (11.7%) | 1440 (19.5%) | 1269 (12.3%) |
| 15 | 1810 (24.7%) | 1461 (23.0%) | 1909 (26.9%) | 1597 (24.7%) | 1455 (25.2%) | 1889 (25.5%) | 2555 (24.8%) |
| 16 | 1911 (26.0%) | 1618 (25.4%) | 1679 (23.7%) | 1628 (25.2%) | 1493 (25.9%) | 1873 (25.3%) | 2708 (26.3%) |
| 17 | 1750 (23.8%) | 1576 (24.8%) | 1707 (24.0%) | 1578 (24.4%) | 1337 (23.2%) | 1697 (23.0%) | 2494 (24.2%) |
| 18+ | 1065 (14.5%) | 1065 (16.7%) | 1158 (16.3%) | 940 (14.5%) | 812 (14.1%) | 494 (6.7%) | 1288 (12.5%) |
| **Median [Q1, Q3]<sup>†</sup>** | **16.6 [15.6, 17.5]** | **16.7 [15.6, 17.7]** | **16.6 [15.5, 17.6]** | **16.5 [15.6, 17.6]** | **16.5 [15.5, 17.5]** | **16.2 [15.2, 17.2]** | **16.5 [15.5, 17.4]** |
| **Mean (SD)<sup>†</sup>** | **16.6 (1.3)** | **16.6 (1.3)** | **16.6 (1.3)** | **16.6 (1.3)** | **16.5 (1.3)** | **16.2 (1.4)** | **16.5 (1.3)** |
| **Female adolescents** |  |  |  |  |  |  |  |
| **Age, years<sup>§</sup>** |  |  |  |  |  |  |  |
| ≤14 | 920 (13.0%) | 695 (10.6%) | 804 (11.6%) | 866 (12.9%) | 743 (12.8%) | 1526 (21.0%) | 1221 (12.8%) |
| 15 | 1750 (24.8%) | 1654 (25.2%) | 1762 (25.5%) | 1633 (24.3%) | 1427 (24.6%) | 1829 (25.1%) | 2468 (25.8%) |
| 16 | 1826 (25.9%) | 1641 (25.0%) | 1797 (26.0%) | 1742 (26.0%) | 1497 (25.8%) | 1794 (24.6%) | 2454 (25.6%) |
| 17 | 1709 (24.2%) | 1623 (24.7%) | 1629 (23.6%) | 1620 (24.1%) | 1408 (24.2%) | 1799 (24.7%) | 2371 (24.8%) |
| 18+ | 850 (12.0%) | 947 (14.4%) | 909 (13.1%) | 850 (12.7%) | 732 (12.6%) | 334 (4.6%) | 1059 (11.1%) |
| **Median [Q1, Q3]<sup>†</sup>** | **16.5 [15.5, 17.5]** | **16.6 [15.6, 17.6]** | **16.5 [15.5, 17.5]** | **16.5 [15.5, 17.5]** | **16.5 [15.5, 17.5]** | **16.2 [15.2, 17.2]** | **16.4 [15.5, 17.5]** |
| **Mean (SD)<sup>†</sup>** | **16.5 (1.2)** | **16.6 (1.3)** | **16.5 (1.2)** | **16.5 (1.2)** | **16.5 (1.3)** | **16.2 (1.4)** | **16.4 (1.2)** |

\* A weight was applied to each student record to adjust for student nonresponse and the distribution of students by grade, gender, and race/ethnicity in each jurisdiction.
<sup>†</sup> Age (decimal places) estimated by multiple imputation separately for each survey year
<sup>§</sup> Participants within missing data on ever had sex (i.e., primary analysis population in **Figure 2**)

Figure 2 shows the weighted age distribution from 2011 to 2023 among students with complete data on age, gender, and sexual intercourse (i.e., ‘ever had sex’ survey question). Corresponding numbers, proportions and averages for gender-specific proportions are provided in **Table 1**, as well as for the overall weighted population (i.e., including those with missing values). From 2011-2019, the age structure in the YRBS national sample showed no significant change (p<.0538). However, compared to other years, the 2021 student population was substantially younger; the percentage of female students aged 14 or younger increased from 13.0% to 21.0% and the percentage of female students aged 18 or older dropped from 12.0% to 4.6%; changes in the age distribution among male students were similar.

**Figure 2.**
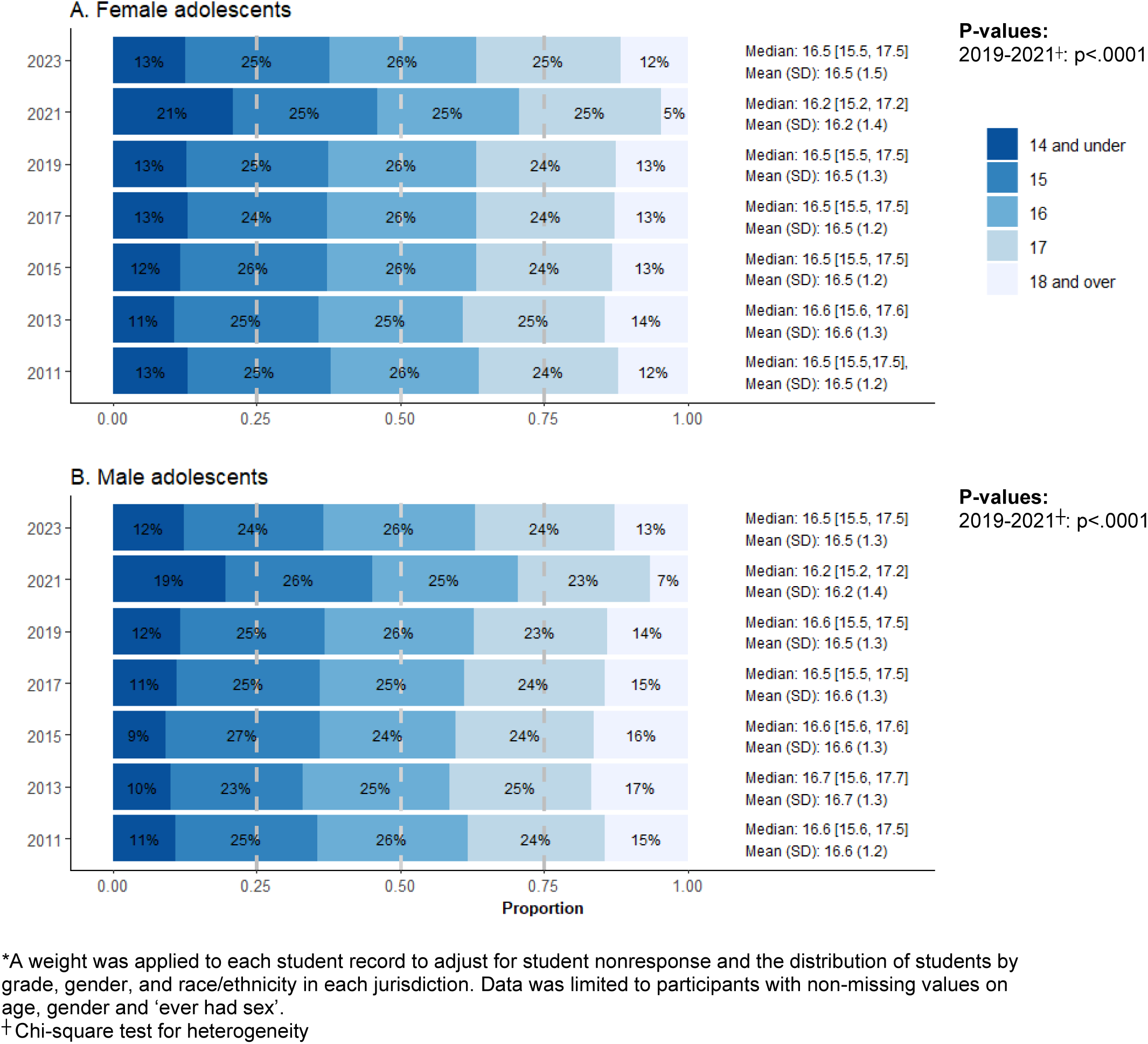
Gender-specific age distribution by study year among national YRBS survey participants*, by study year 2011-2023

Figure 2 also shows mean and median ages. During the years of spring administration (2011-2019, and 2023), the median ages were similar (median=16.5-16.7). However, after moving to Fall administration in 2021, there was an appreciable decline in the median age to 16.2 [15.2, 17.2] among female and male students. The median and mean ages in 2021 were approximately 3-4 months younger compared to the medians during 2011-2019 and 2023 surveys.

Figure 3 compares the unadjusted and age-adjusted percent of students who reported ever having sexual intercourse by study year and sex (exact proportions provided in **Supplementary Table 1**). The most noticeable difference between unadjusted and adjusted rates were observed between 2019 and 2021 for both genders, whereby unadjusted rates appeared to exaggerate the decline in rates of ever reporting having sex. Among female students, prior to adjusting for age, the proportion of female students who ever had sex decreased from 37.6% to 30.6% from 2019 to 2021 or 7.1 percentage points. However, after adjusting the 2021 data to the 2019 age distributions, the percentage who reported ever having sex in 2021 increased to 34.1%; thus, the adjusted change between these years was from 37.6% to 34.1% or 3.5 percentage points. We calculate that 50.4% of the change in ever had sex among female students from 2019 to 2021 was from changes in the age distribution, while the remainder (49.6%) was due to change in adolescent reporting of sexual experience (Figure 4).

**Figure 3.**
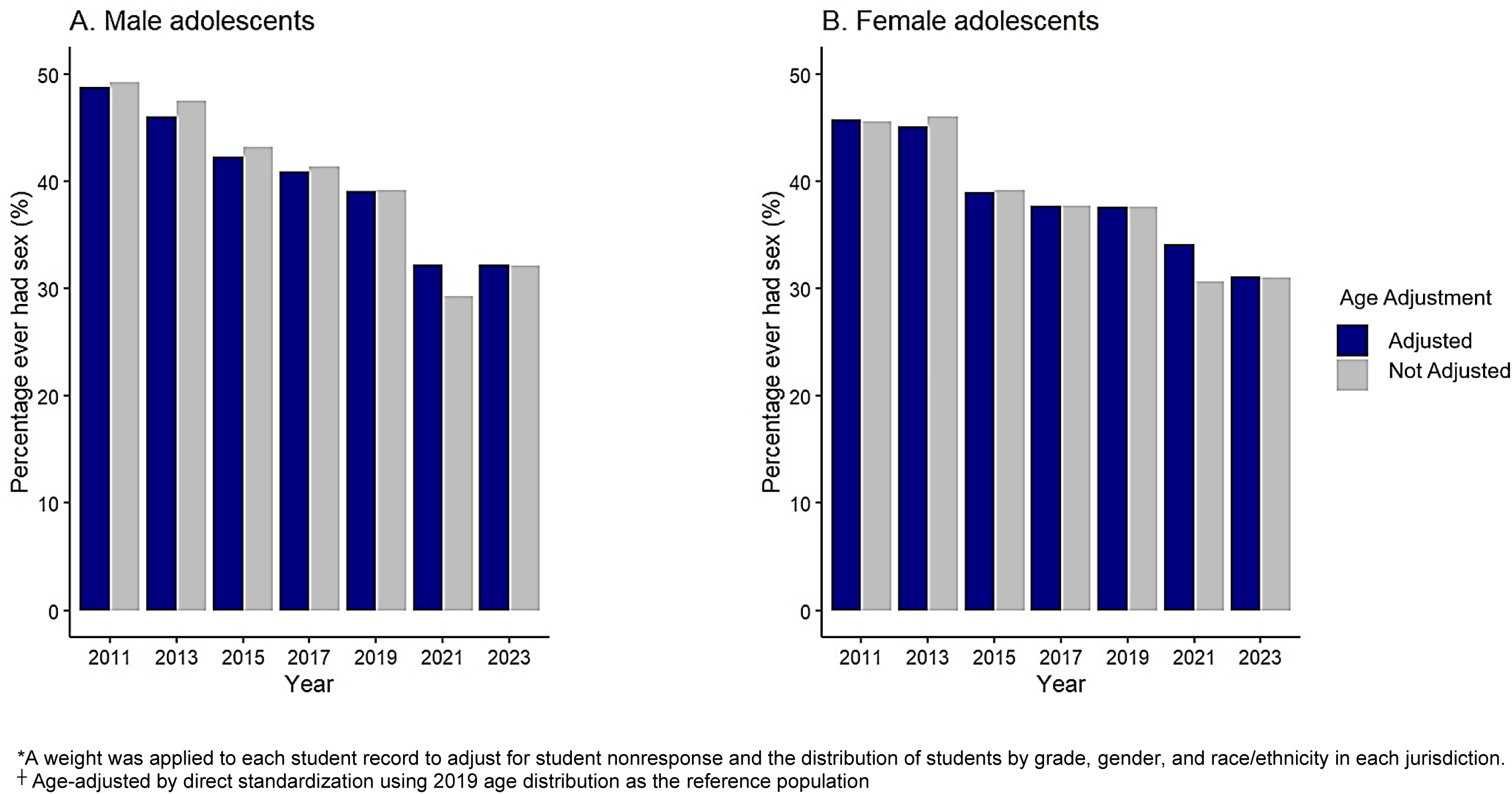
Weighted* percentage of students who ever had sexual intercourse by gender, age-adjusted^┼^ and not age-adjusted, national YRBS, 2011-2023

**Figure 4.**
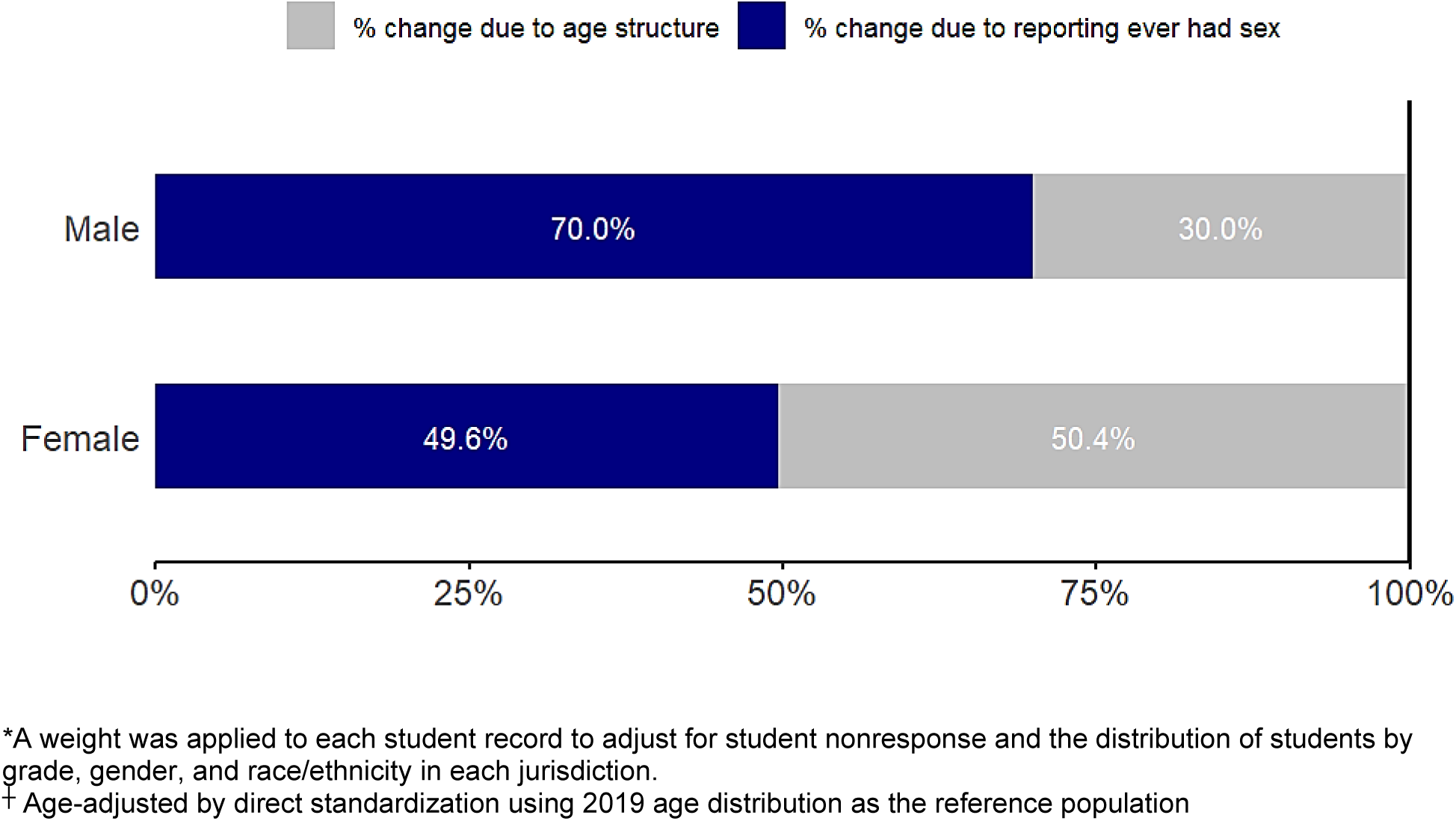
Attribution of change in weighted* percentage of adolescents who ever had sexual intercourse before and after age-adjustment^┼^ by gender, national YRBS 2019-2021

Among male students, in the unadjusted analysis, the proportion who reported ever had sex decreased from 39.1% to 29.3% from 2019 to 2021, or 9.8 percentage points. This is the same change reported by CDC [9]. This change diminished after standardizing the 2021 data to the 2019 age distribution; the adjusted change between these years was from 39.1% (2019) to 32.2% (2021 age-adjusted) or 6.9 percentage points. We calculated that 30% of the change in ever had sex among male students from 2019 to 2021 was due to changes in the age distribution while the majority (70%) was due to change in adolescent reporting of sexual experience.

Finally, to address missing data on ever-had sex, we used data from the *age at first sex* question from 2019 and 2021 YRBS surveys. Incorporating the values from *age at first sex* increased the analytic sample sizes from 10,940 to 11,949 in 2019 and from 11,981 to 15,341 in 2021. The estimated proportion who ever had sex changed very little compared to using only data from the ever-had sex question and the decline from 2019 to 2021 persisted, however, the change in age structure explained a greater proportion of the change in this sensitivity analysis, specifically among female students (See **Supplementary** Figure 2).

## Discussion

The national YRBS has encountered multiple methodological challenges during the period from 2011-2023, including changes in the age structure in 2021, increased missing data on ever-had sex, and declining response rates. These challenges raise serious concerns about the validity of the YRBS data and the validity of trends that have been reported for American high school students. We found a systematic change in the age structure of the YRBS national sample in 2021 compared to the five prior rounds of the national YRBS and the 2023 round. Statistical decomposition estimated that about half of the change in sexual experience among adolescent women between 2019 and 2021 was due to a change in the age distribution; the corresponding estimate for adolescent males was 30%. We also observed a substantial increase over time in the proportion of missing data on ever-had sex, raising concerns about potential bias that could impact estimates. The national YRBS also experienced steady declines in school and student response rates over the 12-year study period. The response rate in 2023 was 35%; such a low response rate also raises the possibility of bias in YRBS estimates of adolescent behaviors.

National YRBS sampling and data collection are presumably influenced by increasing unwillingness by some states to participate fully in the CDC surveillance efforts; this lack of participation is not reported. State YRBS surveys have their own sampling and implementation; however, states which limit or refuse to collect state-level data may also not fully participate in the national survey. To understand participation in the state YRBS, we used the CDC’s online Youth Explorer system^20^; we found that states are increasingly unwilling to ask the ever-had sex question in the state YRBS and this may have influenced data collection in the national YRBS. In 2021, 18 states did not have data on ever had sex. Of these, 6 states did not participate in the YRBS, 4 states did not include any sex questions, and 8 states did not include the ever-had sex question but included other sex questions. By comparison, in 2011, 8 states did not participate, 4 states did not include any sex questions, and 3 states did not include the ‘ever had sex’ question in their state YRBS survey but asked other sex questions.

Declining participation in the YRBS and dropping questions on sex may reflect US culture wars. U.S. schools have experienced intense political and cultural battles about talking about sex^21^. Delivery of sex education to high school students in the United States has declined given political pressures^22,23^. We note that there is little evidence that asking about sex on questionnaires or talking about sex in school-based education increases adolescent sexual activity^24,25^.

Our study examined how a change in the timing of a school survey could affect the age structure of an adolescent sample and the prevalence of sexual experience reported by the surveillance system. Surveillance reports from the YRBS have focused on differences by grade among high school students, with limited attention given to variation by chronological age. Thus, while the YRBS survey sample remains students in grades 9 – 12, high school students are systematically younger in the fall semester compared to the spring. This differential age structure influenced the observed estimates of changes in sexual experience in 2021 compared to earlier years. Beyond age, the social lives and experiences of adolescents are systematically different in the fall and the spring, and these may influence initiation of sexual experience. For example, initiation of sexual intercourse varies seasonally and is more common in the summer months and over the Christmas holiday^26^. Adjusting for age does not adjust for these other differences in adolescent lives.

The COVID-19 pandemic beginning in early 2020 disrupted adolescent schooling and social interactions^22^. The pandemic brought dramatic social and economic changes, including social distancing, stay-at-home requirements, social isolation, barriers to healthcare access and health education, nearly universal school closures, increased engagement with parents or other household members, and growing economic insecurity. These changes may have affected patterns of adolescent initiation of sexual intercourse during the pandemic.

Beyond the issues we examined, the population of US high school students has experienced other historical changes that may influence patterns of sexual behaviors and their surveillance. These include declines in school dropout rates – rates declined from 12.5% in 2005, to 8.3% in 2012 and to 5.8% in 2021. Likewise, changes have occurred in the racial and ethnic composition of the high school population (particularly increases in Hispanic youth) and increases in percentage of foreign-born students^27^.

Data on sexual behavior from the National Survey of Family Growth has been used data to validate estimates for ever had sex in the YRBS - with mixed results^1,3^. Point estimates for ever had sex are much higher on the YRBS compared to the NSFG, although time trends are similar. Data released in December 2024 from the 2022-2023 NSFG also shows plummeting responses rates – from 63.4% in 2017-2019 to 23.4% in 2022-2023. Further work is needed to improve response rates and improve data quality.

## Limitations

As noted above, age adjustment may not remove the systematic bias in a student sample as high school students may be different in spring and fall on factors other than age. Age-adjustment cannot control for these unmeasured differences over the school year. All data in the YRBS are self-reported and adolescent willingness to self-report sensitive behaviors can be influenced by multiple factors including adolescent sense of privacy and the mode of data collection, which could change over time. Our analyses of data on national YRBS cannot directly answer questions about the validity of the data in the face of declining response rates.

## Implications

Consistency in data collection for public health surveillance systems is essential. In many ways, the YRBS has achieved strong consistency in data collection and has demonstrated good reliability since its inception in 1991. However, changes to the timing of a school-based survey can influence age structure, induce seasonal factors, and alter estimates of health behaviors. Increases in missing data on the ever-had sex questions over time and declining response rates over time also raise growing concerns about the validity of the YRBS data. School-based surveillance systems like the YRBS should strive for consistency in the timing of data collection and consider methods to increase state, school, and student participation. School-based surveillance systems should report changes in age distribution and consider adjustment for age in reporting behavioral data, although the latter will not fix other differences such as seasonal factors. The 2021 YRBS data should not be used by researchers to estimate changes in sexual experience and other adolescent behaviors in comparison to prior survey years without appropriate age adjustments or explanations. The YRBS online system should add online functionality to adjust trend data for age and provide warnings to data users about the age structure in the 2021 survey.

The US is experiencing a crisis in participation in national surveys^6^ including the national YRBS. While declining responses rates is not new, the crisis is now existential. A 2016 report^6^ on this issue suggests a number of technical fixes such as increasing compensation for research participation. But in the face of plummeting response rates, additional approaches are needed. Adolescent health is a concern across the political spectrum. Both conservative and progressive parents share concerns about adolescent health and would benefit from data on trends in adolescent health behaviors that can be used to assess health needs, build public health programs, and generate community buy-in to such programs. The solution to this crisis must include community, parent, and adolescent mobilization. Leadership from national and local public health policy makers will be essential.

## Abbreviations

(YRBS): Youth Risk Behavior Survey
(CDC): Centers for Disease Control and Prevention
(DOE): Department of Education

## Data Availability

All data produced in the present study are available online and upon request to the authors.

