## Supplementary Appendix for "Age Bias, Missing Data, and Declining Response Rates in the National Youth Risk Behavior Survey and Their Influence on Estimates of Trends in Adolescent Sexual Experience, 2011-2023"

**Supplementary table 1**. Gender-specific crude and weighted proportion of national YRBS respondents reporting 'ever had sex' with and without age-adjustment*, 2011-2023

| Study year | Row total | % reporting ever had sex without age adjustment ┼ | % reporting ever had sex with age adjustment § |
|---|---|---|---|
| **Females** | | | |
| 2011 | 7056 | 45.6% | 45.8% |
| 2013 | 6560 | 46.0% | 45.1% |
| 2015 | 6901 | 39.2% | 39.0% |
| 2017 | 6711 | 37.7% | 37.7% |
| 2019 | 5806 | 37.6% | 37.6% |
| 2021 | 7281 | 30.6% | 34.1% |
| 2023 | 8243 | 31.0% | 31.1% |
| **Males** | | | |
| 2011 | 7339 | 49.2% | 48.8% |
| 2013 | 6360 | 47.5% | 46.0% |
| 2015 | 7099 | 43.2% | 42.3% |
| 2017 | 6465 | 41.4% | 40.9% |
| 2019 | 5775 | 39.2% | 39.1% |
| 2021 | 7394 | 29.3% | 32.2% |
| 2023 | 8820 | 32.1% | 32.2% |

* Data included in **Figure 3**. A weight was applied to each student record to adjust for student nonresponse and the distribution of students by grade, gender, and race/ethnicity in each jurisdiction.

┼ YRBS data reported in 2021 report for 'ever had sex'

§ Direct standardization using 2019 age distribution as the reference population

**Supplementary figure 1.** Study population of YRBS population included in the primary and sensitivity analyses to assess sexual risk behaviors, 2011-2023

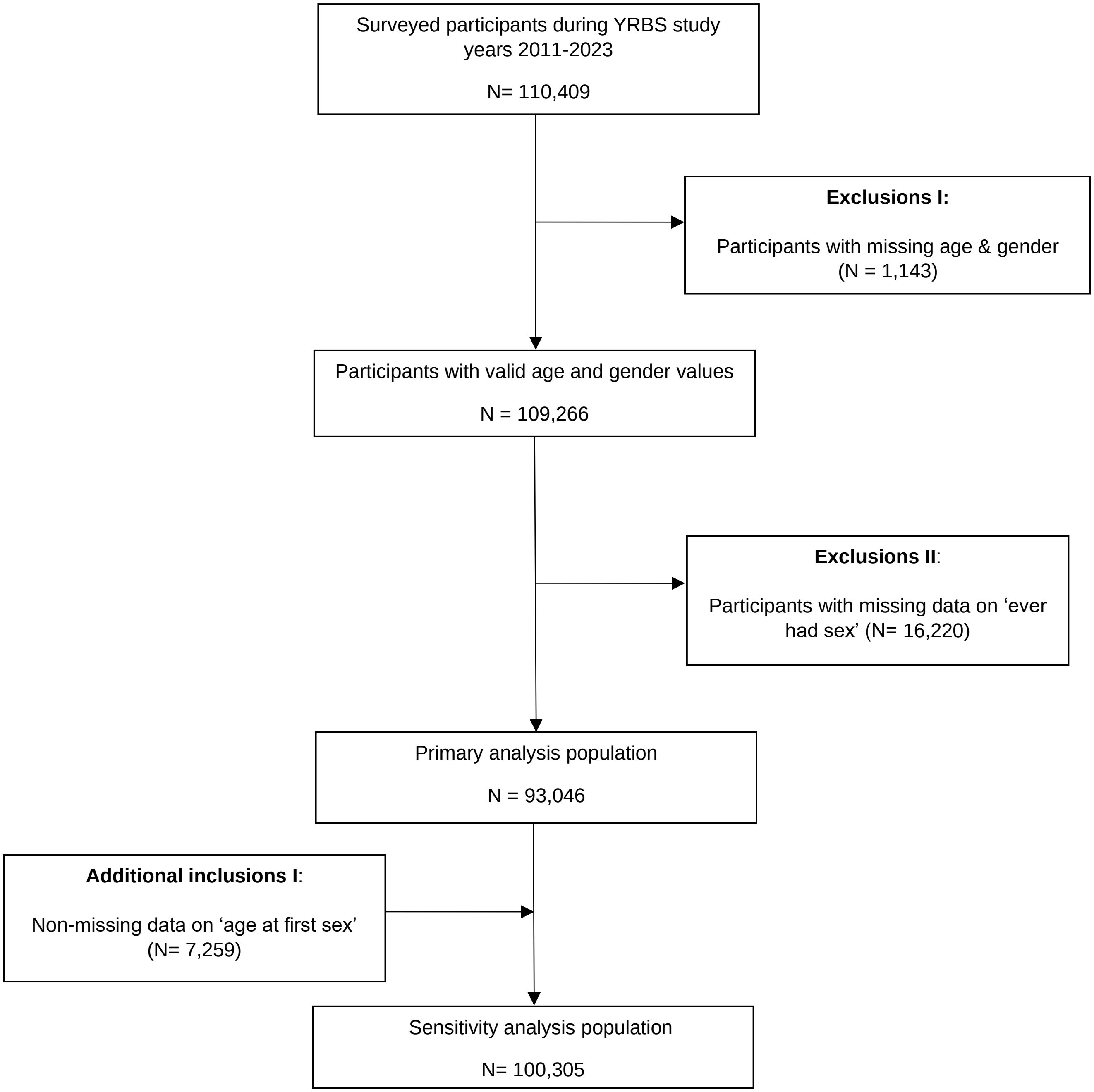

**Supplementary figure 2.** Sensitivity analysis demonstrating the attribution of change in ever had sexual intercourse, before and after age-adjustment┼ by gender, national YRBS, 2019-2021

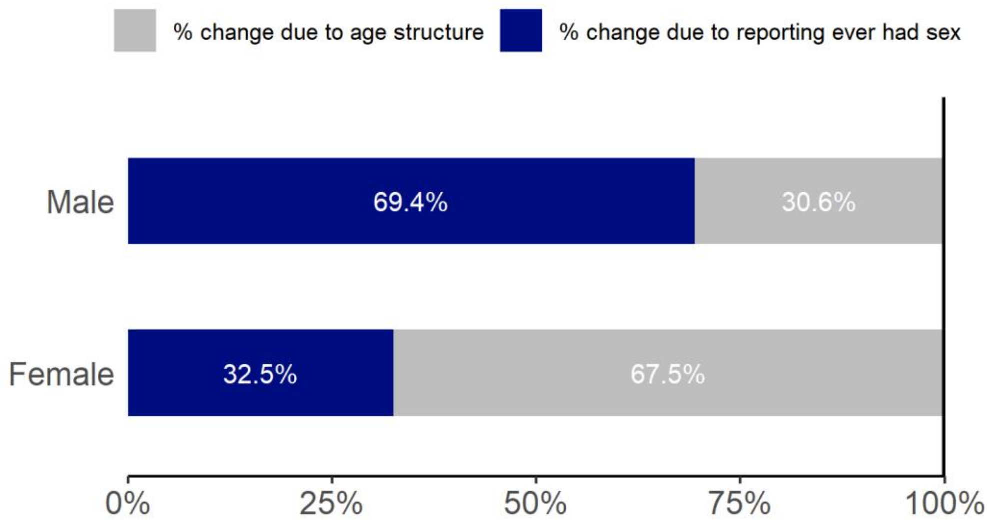


When ‘ever has sex” was missing, age at first sex was used to impute the missing values.
* A weight was applied to each student record to adjust for student nonresponse and the distribution of students by grade, gender, and race/ethnicity in each jurisdiction.
┼ Age-adjusted by direct standardization using 2019 age distribution as the reference population
